# PHIVE: A Physics-Informed Variational Encoder Enables Rapid Spectral Fitting of Brain Metabolite Mapping at 7T

**DOI:** 10.1101/2025.01.02.25319930

**Authors:** Amirmohammad Shamaei, Eva Niess, Lukas Hingerl, Bernhard Strasser, Aaron Osburg, Korbinian Eckstein, Wolfgang Bogner, Stanislav Motyka

**Affiliations:** Electrical and Software Engineering, University of Calgary, Calgary, AB, Canada; Hotchkiss Brain Institute, University of Calgary, Calgary, AB, Canada; Department of Biomedical Imaging and Image-guided Therapy, Radiology and Nuclear Medicine, Medical University of Vienna, Vienna, Austria; School of Electrical Engineering and Computer Science, The University of Queensland, St Lucia, Australia; Christian Doppler Laboratory for MR Imaging Biomarker Development, Vienna, Austria

**Keywords:** Deep learning, MRSI, Whole brain, Quantification

## Abstract

Magnetic Resonance Spectroscopic Imaging (MRSI) enables non-invasive mapping of brain metabolite concentrations but remains computationally intensive and challenging due to a low signal-to-noise ratio (SNR) and overlapping spectral features. Traditional spectral fitting methods, such as LCModel, are time-consuming and often lack comprehensive uncertainty quantification. In this study, we propose Physics-Informed Variational Encoder (PHIVE), a novel deep learning framework that integrates physics-based priors into a variational autoencoder architecture for rapid and accurate metabolite quantification. PHIVE enables simultaneous estimation of metabolite concentrations and uncertainty metrics, including Cramér-Rao Lower Bound (CRLB), aleatoric, and epistemic uncertainties.

PHIVE was evaluated on whole-brain MRSI data from 7T acquisitions of healthy controls. The method achieved comparable accuracy to LCModel for key metabolites, such as Total N-acetylaspartate (tNAA), Glutamate-Glutamine complex (Glx), and Myo-inositol (mIns) while demonstrating a six-order magnitude reduction in computational time (6 ms per dataset). Uncertainty quantification highlighted PHIVE’s robustness in regions with low SNR. Additionally, a conditional baseline modeling approach was introduced, enabling dynamic flexibility in spectral baseline estimation during inference time.

These results suggest that PHIVE offers a fast, reliable, and interpretable solution for high-resolution metabolite quantification, paving the way for real-time MRSI applications in clinical and research settings. Future work will focus on expanding its validation across diverse datasets and investigating its utility in longitudinal and multicenter studies.

## 1. Introduction

MRSI is a powerful technique that maps the spatial distribution of metabolite concentrations in various regions of the body (de Graaf (2019)). Whole-brain MRSI with 3 mm isotropic resolution can be achieved by applying spatial-spectral encoding in a clinically attractive time of 15 minutes (Hingerl et al. (2020); Bogner et al. (2021)). However, the reconstructed MRSI data are a 3D matrix of spectra, which are not directly interpretable and must be fitted by parametric models to obtain concentration estimates or ratios of chemical compounds within these spectra (Near (2014); Provencher (2001)). The spectral fitting of MRSI data poses several challenges, primarily due to the low SNR and the presence of nuisance signals such as residual water peaks and strong lipid signals relative to the brain metabolites. Furthermore, MRSI generates hundreds of thousands of spectra that require fitting, making the process computationally intensive and time-consuming (Maudsley et al. (2021)). Parametric fitting methods, such as those proposed by Provencher et al. (Provencher (2001)), were initially developed for quantifying single voxel spectra. However, when applied to whole-brain MRSI, the number of spectra to be quantified increases by orders of magnitude. The uncertainty in the model fit parameter estimation is commonly addressed using CRLB since direct access to the uncertainty is impossible without repeated measurements (Near (2014)). The CRLB states that the variance of any unbiased estimator is lower bounded by the inverse of the Fisher information. Traditional approaches are generally slow and computationally demanding, which makes them impractical for clinical use. Deep learning has emerged as a powerful tool in various aspects of magnetic resonance imaging (MRI) and MRSI (Lundervold and Lundervold (2019)). These techniques have shown promise in addressing the challenges associated with traditional spectral analysis methods, offering faster and more efficient solutions. However, conventional neural networks lack an understanding of the underlying physical principles governing the observable MRS data, which leads to reduced reliability in metabolite quantification, potential overfitting to noise patterns, and inability to generalize well to data acquired under different experimental conditions (Shamaei et al. (2023)). These limitations are particularly problematic in clinical applications where accurate and robust metabolite quantification is crucial for diagnosis and treatment monitoring. Moreover, most of the deep learning approaches currently studied in the MRS field are limited to supervised learning, where ground truth knowledge is required. In reality, acquiring hundreds of spectra with various ground truth values is impractical for supervised training. Previous works by Gurbani et al. (Gurbani et al. (2019)) and Shamaei et al. (Shamaei et al. (2023)) presented unsupervised physics-informed neural network (Physics-Informed Neural Network (PINN)) architectures that incorporate parametric analyses into the network structure, employing an autoencoder (AE) module Goodfellow et al. (2016) to map high-level features of the data to smaller subspaces. Building upon these advancements, we propose the following contributions:

1. Propose a novel deep learning framework called (PHIVE) for high-resolution metabolite quantification with simultaneous CRLB estimation.
2. Incorporate a variational autoencoder within the network architecture to model the latent space probabilistically, accounting for the estimation of aleatoric and epistemic uncertainties in the latent representations and propagating them to the final metabolite concentration estimates.
3. Evaluate the proposed PHIVE framework using highresolution whole-brain MRSI data of healthy volunteers obtained at 7T, aiming to achieve a substantial gain in quantification speed compared to established approaches.

## 2. Methods

### 2.1. In vivo data

All *in vivo* MRSI data were acquired at a 7T Magnetom+ MR Scanner (Siemens Healthineers, Erlangen, Germany) with a 32-channel head coil (Nova Medical, Wilmington, MA). Data from 12 healthy subjects (4 male, and 8 female, age 34.4+-6.7) were used in this study.

MRSI data were measured with an echo-less slab-selective 3D MRSI sequence with concentric ring trajectory based spatial-spectral encoding of a spherical k-space (Hingerl et al. (2020)): FOV:220×220×133 mm3, matrix size: 64×64×33, resolution 3.4×3.4×4.0mm3, TA: 8:30 min, TR: 320 ms, acquisition delay 1.3 ms, excitation flip angle 34 deg., bandwidth: 2778 Hz, with three variable temporal interleaves and WET water suppression.

(Gold) standard LCModel spectral processing Provencher (2001) for comparison was performed in LCModel (V6.3-1, LCModel Inc, Oakville, ONT, Canada) with a simulated basis set that included 17 brain metabolites and a measured macro-molecular background over an evaluation range 1.8-3.88 ppm. Prior to spectral quantification, raw data were processed using in-house developed pipeline written in Matlab and Bash, which included iMUSICAL coil combination, k-space reconstruction with in-plane convolution gridding, spatial Hamming filtering, channel-wise noise decorrelation, and lipid signal removal using L2-regularization.

Our study included a total of twelve subjects, with eight healthy subjects used for training of our model (122473 spectra) and four for testing (39051 spectra).

### 2.2. Neural networks

#### 2.2.1. Variational Inference

Variational inference is formulated as the issue of identifying a model distribution *q*(**z**) that closely approximates the true posterior *p*(**z**|**x**) as follows:

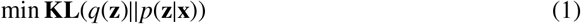

where the Kullback-Leibler divergence (**KL**) is a measure of the difference between two probability distributions. The **KL** may then be modified in order to establish a lower bound on the intractable marginal probability *p*(**x**). Evidence Lower Bound (ELBO) derived from Eq. 1 is given by

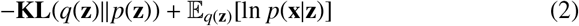

The first term represents the reconstruction likelihood, and the second term ensures that our learned distribution *q*(**z**) is similar to the true prior distribution *p*(**z**). Eq. 1 can be minimized by maximizing Eq. 2 (ELBO). In the case of latent variable models, the distribution of the model directly relies on **x**, i.e., *q*(**z**|**x**). The encoder and decoder are trained jointly using the ELBO, which regularizes the data likelihood under the approximate posterior with the **KL** between the true posterior and the suitable parametric posterior for *p*(**z**) and *q*(**z**|**x**). The second term in Eq. 2 is stochastic with respect to **z** (the parameters of linear combination model). Because **z** is a random variable and taking the derivative of a random variable is not possible, neural networks cannot be trained using the backpropagation algorithm. However, **z** can be expressed using a differentiable transformation, i.e., **z** = *g*_*φ*_(ϵ, *x*) where *ϵ* is an auxiliary variable with independent marginal *p*(*ϵ*), and *g*_*φ*_(.) is some vector-valued function parameterized by *φ*. In the case of univariate Gaussian in which **z** = *p*(**z**|**x**) = 𝒩(*µ, σ*), a valid reparameterization is **z** = *µ* + *σϵ* where *ϵ* is an auxiliary noise variable *ϵ* ∼ 𝒩(0, 1).

#### 2.2.2. Physics-informed Variational Encoder

A PHIVE can be designed by implementing *q*(**z**|**x**) and *p*(**x**|**z**) as neural networks and physics-based decoder, respectively. A continuous latent variable model is designed to encode the *n*-dimensional input vector (**x** ∈ ℝ^*n*^) to the *n*^′^ -dimensional latent vector of model’s parameters (**z** ∈ ℝ(*n*^′^), where *n*^′^ < *n*). The model consists of a learnable encoding model *q*(**z**|**x**) and a physics-based decoding model *p*(**x**|**z**) given *p*(**z**). *q*(**z**|**x**) can be computed as follows:

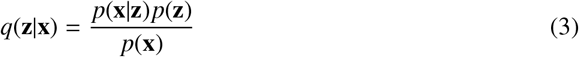

where *p*(**x**) = *∫ p*(**x**|**z**)*p*(**z**)d**z**. Typically, the distribution *p*(**x**) reveals to be intractable. However, we may estimate its value via variational inference.

Our proposed method (PHIVE) is a deep learning model designed to learn low-dimensional latent representations of high-dimensional systems governed by known physical laws. As depicted in Figure 1 PHIVE consists of a convolutional encoder that maps input data into a latent space, which is then used by a physics-informed decoder to reconstruct the original data.

**Fig. 1.**
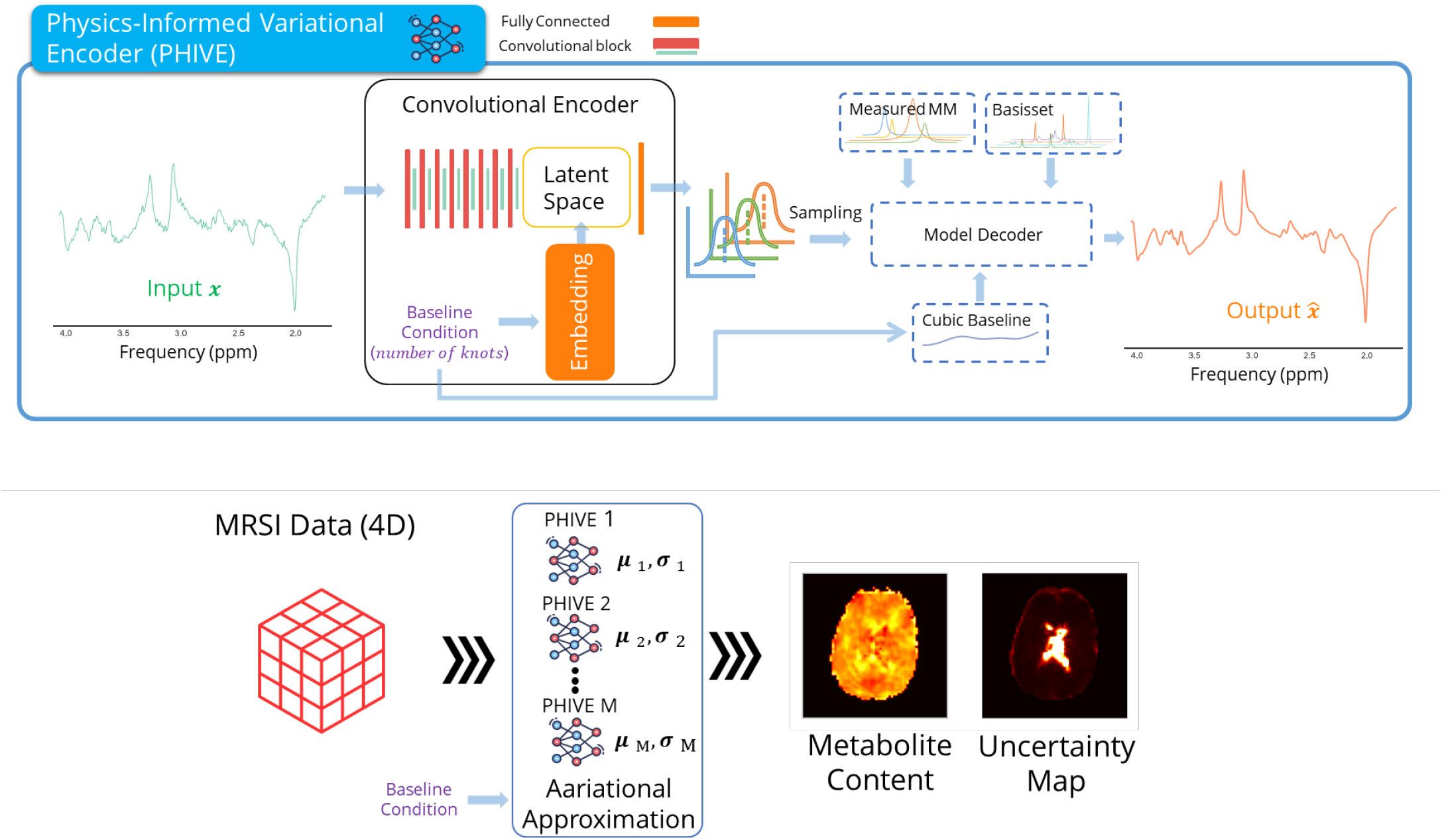
(A) Overview of the PHIVE architecture. The input data (*x*) is passed through a convolutional encoder to obtain a low-dimensional latent representation. The latent space is constrained by the governing physical equations. The decoder samples from this constrained latent space and reconstructs the original data (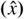). The model is trained by minimizing the difference between *x* and 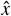 while ensuring the latent space follows the physical constraints. (B) Application of the trained PHIVE model during inference time for quantifying metabolite content and generating uncertainty maps. The input data is passed through the encoder to obtain the latent representation, which is then used by the decoder to reconstruct the metabolite content map and the associated uncertainty map.

The encoder consisted of dropout layers (Schmidhuber (2015)), normalization layers, convolutional layers (Schmidhuber (2015)), and LeakyReLU activation units. A fully connected layer was used to compute four parameters—amplitude, frequency, phase, and damping—which served as inputs to the model decoder from the latent space. Additionally, a Softplus function (Goodfellow et al. (2016)) was applied to the estimated amplitudes to ensure non-negative values for the relative metabolite concentrations (amplitudes).

The model is trained by minimizing the difference between the input and reconstructed data while ensuring the latent space adheres to the governing physical equations. This physics-informed constraint allows PHIVE to learn meaningful and interpretable latent representations that capture the underlying dynamics of the system. Training our proposed network is an unsupervised learning task that does not require ground truth values. It minimizes the differences between the original input and the consequent reconstructions. In each iteration step of training, the parameters of the encoders were adjusted according to the gradient of the loss function with respect to the given parameters of the model (*A*_*M*_, Δ*α*, Δ *f*, and Δ*θ*). Assuming *p*(*z*) = *N*(0, *I*), the mathematical representation of the loss function of training can be written as follows:

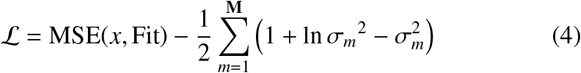

where MSE is the mean square error, *x* and Fit are the *n*-dimensional input vector (*x* ∈ ℝ^*n*^) of the encoder and the *n*-dimensional output vector (Fit ∈ ℝ^*n*^) of the model-decoder, respectively, and *σ*_*m*_ is the variational standard deviation of the *m*-th metabolite amplitudes. Gradients were computed with the help of PyTorch using its automatic gradient computation (**?**), which is a reverse automatic differentiation system. The input and outputs of the PHIVE were the real parts of signals in the frequency domain in the range of 1.8 ppm to 3.88 ppm. The concentration of metabolites is quantified using the signal intensity of each metabolite’s basis set. The measurements are expressed in arbitrary units for relative quantification, enabling straightforward comparisons between samples.

### 2.3. Metrics for uncertainty estimation

Lakshminarayanan et al. (Lakshminarayanan et al. (2016)) empirically demonstrated that the deep ensembles method is a potential technique for enhancing the accuracy, quantifying uncertainty, and out-of-distribution robustness of DL models.

We utilized a randomization-based ensemble model combination in which five proposed PHIVEs with randomly initialized parameters were trained independently using the whole training dataset. Interestingly, Bayesian neural networks can be viewed as an ensemble of randomly initialized neural networks that averages the estimates. Thus, the variance of estimates of the networks is the sum of aleatory and epistemic uncertainty. Since the encoder of our proposed method can estimate a mean and variance for each metabolite, the aleatory and epistemic uncertainty for each metabolite can be quantified as follows:

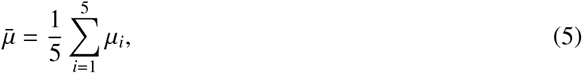

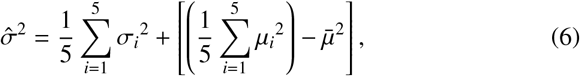

where *µ* and *σ* are the variational mean and standard deviation of the metabolite amplitude. The first term in variance stands for aleatory uncertainty, and the second term stands for epistemic uncertainty (Kendall and Gal (2017)).

#### 2.3.1. Baseline construction

A linear combination of natural cubic spline basis functions is utilized to construct the smooth baseline. The spline function is differentiable and the coefficients are obtained from the encoder. This allows the baseline to be optimized as part of the model. There are 8 knots, with 2 knots located at the edges of the fit range and 2 additional knots outside the fit range.

The default knot spacing is increased to 0.36 ppm compared to 0.15 ppm in LCModel (Provencher (2001)). This prevents an overly flexible baseline without using as much regularization as LCModel (Oeltzschner et al. (2020)).

#### 2.3.2. Conditional Baseline

The flexibility of the baseline is a critical factor in accurate metabolite quantification. As highlighted in Simicic et al. (2021), a very stiff baseline in combination with a single Macromolecule (MM) spectrum can lead to deviations in metabolite concentrations.

To address this issue while avoiding the time-consuming process of training multiple networks with different baseline stiffness levels, we propose a conditional baseline approach. In this method, we condition the output of the convolutional neural network (*LS*) using the desired stiffness level.

During training, for each batch, we randomly select an integer value (*c*) from a range of 0 to 3, which represents the stiffness level (where lower values correspond to increased rigidity and higher values impose greater flexibility). This integer is then embedded into a vector representation (*E*(*c*)). The output of the Convolutional Neural Network (CNN) encoder is multiplied by the feature map with the embedded vector before mapping the model parameters (*E*(*c*) × *LS*). Additionally, *c* is corresponded to the number of knots in the baseline (0 → 3, 1 → 6, 2 → 9, 3 → 12). This allows the network to learn to generate baselines with different levels of flexibility based on the input stiffness condition.

During inference, the model takes an integer value from the same range (0 to 3) as input, representing the desired stiffness level. The model then fits the input signal conditioned on this stiffness class. Higher values correspond to stiffer baselines, while lower values allow for more flexibility.

By incorporating this conditional baseline approach, our model can dynamically adjust the baseline flexibility based on the input stiffness level. This eliminates the need for training multiple networks with different stiffness settings and allows for more efficient and adaptable baseline modeling.

#### 2.3.3. Implementation details and training strategy

All steps were run on a computer with a 13th Gen Intel(R) Core(TM) i9-13900HX 2.20 GHz processor and one graphics processing unit (NVIDIA GeForce RTX 4080 16GB). The DAE was implemented in Python with the help of the Pytorch lightning interface (Paszke et al. (2019); Lightning (2020)).

All training was performed using MSE loss and an Adam optimizer (Kingma and Ba (2015)) with a batch size of 128, a learning rate of 0.001, and 200 epochs.

An early-stopping strategy (Paszke et al. (2019)) was performed by monitoring the MSE of the validation subset at the end of every epoch and stopping the training when no improvement was observed in 20 epochs.

Due to phenomena known as **KL** disappearing, training variational autoencoders is often regarded as being extremely challenging (Bowman et al. (2016)). It has been shown (Bowman et al. (2016)) that adding a variable weight to the **KL** term in the cost function at training time would mitigate this problem. We set that weight to 0 at the beginning of training so that the model can learn to encode as much information as possible in latent space. Then, using a sigmoid annealing schedule, we gradually raised this weight as training advanced (Bowman et al. (2016)).

## 3. Data Analysis

In this study, Coefficient of Variation (CV) is used to assess the variability of metabolite measurements relative to the mean value. It is calculated as the ratio of the standard deviation to the mean, expressed as a percentage. A lower CV indicates higher precision in metabolite estimations. To compare the performance of the proposed PHIVE method and the gold standard LCModel, Bland-Altman plots are generated for six key metabolite ratios: tNAA/Total Creatine (tCr), Glx/tCr, Total Choline (tCho)/tCr, mIns/tCr, Taurine (Tau)/tCr, and Glutathione (GSH)/tCr. Additionally, Pearson correlation coefficients (*R*^2^) are calculated to quantify the strength of the linear relationship between the measurements obtained from the two methods. A paired t-test is used to compare the means of metabolite concentrations obtained using PHIVE and LCModel. Statistical significance is determined using a significance level of 0.05 (*p* < 0.05).

## 4. Results

The spectral fitting via PHIVE during the testing phase took 6 milliseconds per whole-brain MRSI dataset, which represents a reduction in computational time by six orders of magnitude compared to our gold standard approach (i.e., LCmodel), which takes 2 hours for the same amount of spectra.

The PHIVE and LCModel methods were applied to test the same datasets to assess their ability to quantify the relative concentrations. Figure 2 presents a visual comparison of the metabolite maps generated by LCModel and PHIVE, along with the difference maps highlighting the discrepancies between the two methods. The metabolite maps produced by PHIVE exhibit a high degree of similarity to those generated by LCModel, indicating that our method can quantify metabolite relative concentrations with comparable accuracy. The difference maps reveal the variations between LCModel and PHIVE across all six metabolites. Notably, the tCho/tCr and mIns/tCr maps show slightly larger differences compared to the other metabolites, with PHIVE estimating higher relative concentrations in some regions compared to LCModel.

**Fig. 2.**
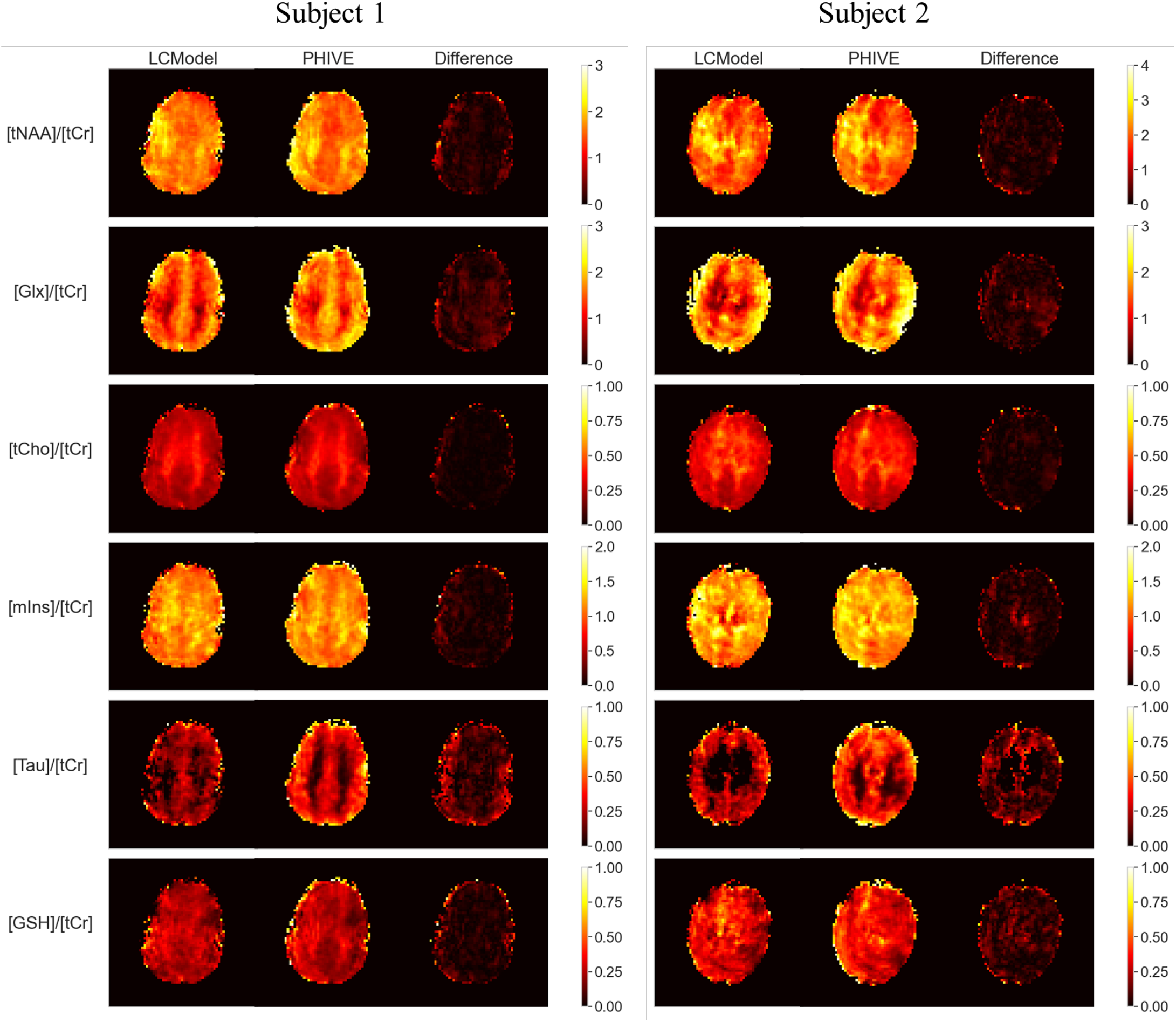
Comparison of metabolite maps generated by LCModel and PHIVE for four key metabolites: tNAA/tCr, Glx/tCr, tCho/tCr, mIns/tCr, Tau/tCr, and GSH/tCr. The first column shows the maps produced by LCModel, the second column displays the maps generated by PHIVE, and the third column presents the difference maps between the two methods. The color scale represents the metabolite relative concentrations, with higher values indicated by warmer colors. The difference maps highlight the discrepancies between LCModel and PHIVE, with the color scale showing the magnitude of the differences.

Figure 3 illustrates the Bland-Altman plots for six key metabolites: tNAA/tCr, Glx/tCr, tCho/tCr, mIns/tCr, Tau/tCr, and GSH/tCr. The correlation plots reveal strong positive correlations between the two measurements for all six metabolites. The *R*^2^ range from 0.27 to 0.86, indicating a high degree of agreement between the two fitting methods. The NAA/tCr and Glx/tCr metabolites exhibit the strongest correlations, with maximum *R*^2^ values of 0.70 and 0.86, respectively. The tCho/tCr, mIns/tCr, Tau/tCr, and GSH/tCr metabolites also show substantial correlations, with maximum R^²^ values of 0.64, 0.62, 0.60, and 0.48, respectively.

**Fig. 3.**
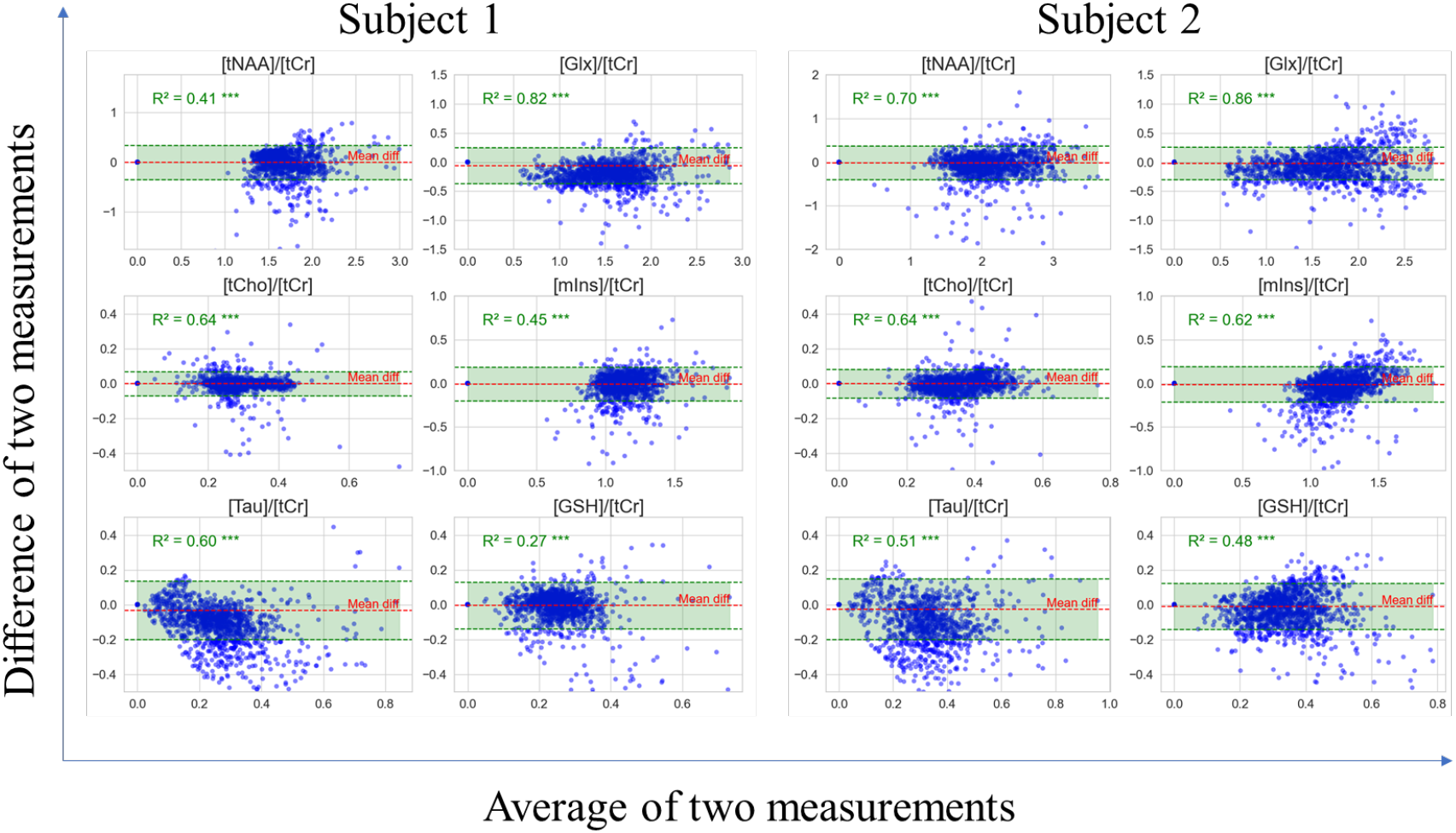
Correlation analysis of metabolite concentrations obtained from two repeated measurements using the PHIVE method. The Bland-Altman plots display the relationship between the first and second measurements for six key metabolite ratios: tNAA/tCr, Glx/tCr, tCho/tCr, mIns/tCr, Tau/tCr, and GSH/tCr. *R*^2^ is provided for each metabolite, indicating the strength of the linear relationship between the two measurements. The diagonal line represents perfect agreement between the measurements. The mean difference (Mean diff) between the two measurements is also shown for each metabolite. The green area indicates the 95% limits of agreement.

The results presented in Table 1 compare the performance of LCModel and PHIVE in estimating metabolite relative concentrations for four test subjects. The table provides the mean ± standard deviation and CV for each metabolite (Glx, mIns, tCho, and tNAA) estimated by both methods. Across all subjects, PHIVE consistently demonstrates lower CVs compared to LCModel for the majority of metabolites. For Glx, PHIVE achieves CVs ranging from 22.10% to 24.10%, while LCModel’s CVs range from 24.17% to 28.70%. Similarly, for mIns, PHIVE’s CVs are between 11.15% and 13.47%, whereas LCModel’s CVs fall between 15.28% and 16.02%. These results highlight PHIVE’s improved precision in estimating Glx and mIns relative concentrations compared to LCModel. The CVs for tCho are comparable between the two methods, with PHIVE slightly outperforming LCModel in most cases. For tNAA, PHIVE demonstrates lower CVs for subjects 2 and 3, while LCModel shows marginally better precision for subjects 1 and 4.

**Table 1.** Mean ± standard deviation and CV for metabolite relative concentrations estimated by LCModel and PHIVE for four subjects.

| Subject | [Metabolite]/[tCr] | Variable | Mean ± Std | CV (%) |
| --- | --- | --- | --- | --- |
| Subject 1 | Glx | LCModel | 1.454 ± 0.388 | 26.66 |
| | | PHIVE | 1.664 ± 0.368 | 22.10 |
| | Ins | LCModel | 1.128 ± 0.181 | 16.02 |
| | | PHIVE | 1.152 ± 0.155 | 13.47 |
| | tCho | LCModel | 0.279 ± 0.069 | 24.64 |
| | | PHIVE | 0.287 ± 0.079 | 27.52 |
| | tNAA | LCModel | 1.666 ± 0.275 | 16.52 |
| | | PHIVE | 1.693 ± 0.279 | 16.48 |
| Subject 2 | Glx | LCModel | 1.467 ± 0.355 | 24.17 |
| | | PHIVE | 1.668 ± 0.380 | 22.79 |
| | Ins | LCModel | 1.155 ± 0.182 | 15.77 |
| | | PHIVE | 1.185 ± 0.159 | 13.45 |
| | tCho | LCModel | 0.308 ± 0.076 | 24.73 |
| | | PHIVE | 0.311 ± 0.077 | 24.64 |
| | tNAA | LCModel | 1.774 ± 0.274 | 15.47 |
| | | PHIVE | 1.912 ± 0.258 | 13.51 |
| Subject 3 | Glx | LCModel | 1.632 ± 0.441 | 27.01 |
| | | PHIVE | 1.783 ± 0.430 | 24.10 |
| | Ins | LCModel | 1.109 ± 0.169 | 15.28 |
| | | PHIVE | 1.193 ± 0.133 | 11.15 |
| | tCho | LCModel | 0.314 ± 0.075 | 23.75 |
| | | PHIVE | 0.328 ± 0.070 | 21.35 |
| | tNAA | LCModel | 1.978 ± 0.341 | 17.25 |
| | | PHIVE | 2.069 ± 0.303 | 14.63 |
| Subject 4 | Glx | LCModel | 1.399 ± 0.401 | 28.70 |
| | | PHIVE | 1.626 ± 0.360 | 22.12 |
| | Ins | LCModel | 1.182 ± 0.186 | 15.74 |
| | | PHIVE | 1.179 ± 0.144 | 12.21 |
| | tCho | LCModel | 0.307 ± 0.067 | 21.81 |
| | | PHIVE | 0.318 ± 0.067 | 21.16 |
| | tNAA | LCModel | 1.703 ± 0.255 | 14.99 |
| | | PHIVE | 1.719 ± 0.278 | 16.18 |

Figure 4 presents a comparison of relative metabolite concentrations obtained using LCModel and our proposed PHIVE method. The relative concentrations of six key metabolites—tNAA, Glx, tCho, mIns, Tau, and GSH—were analyzed. For NAA, PHIVE yields similar relative concentrations to LCModel, with no statistically significant differences observed (*p* < 0.05). In the case of Glx, PHIVE shows a slightly higher relative concentration compared to LCModel, but the difference is not statistically significant (*p* < 0.05). For tCho and Tau, PHIVE demonstrates no statistically significant differences in relative concentrations compared to LCModel (*p* < 0.05). For mIns, PHIVE results in significantly higher relative concentrations compared to LCModel (*p* < 0.05). The relative concentrations of GSH are also similar between the two methods, with no statistically significant differences observed.

**Fig. 4.**
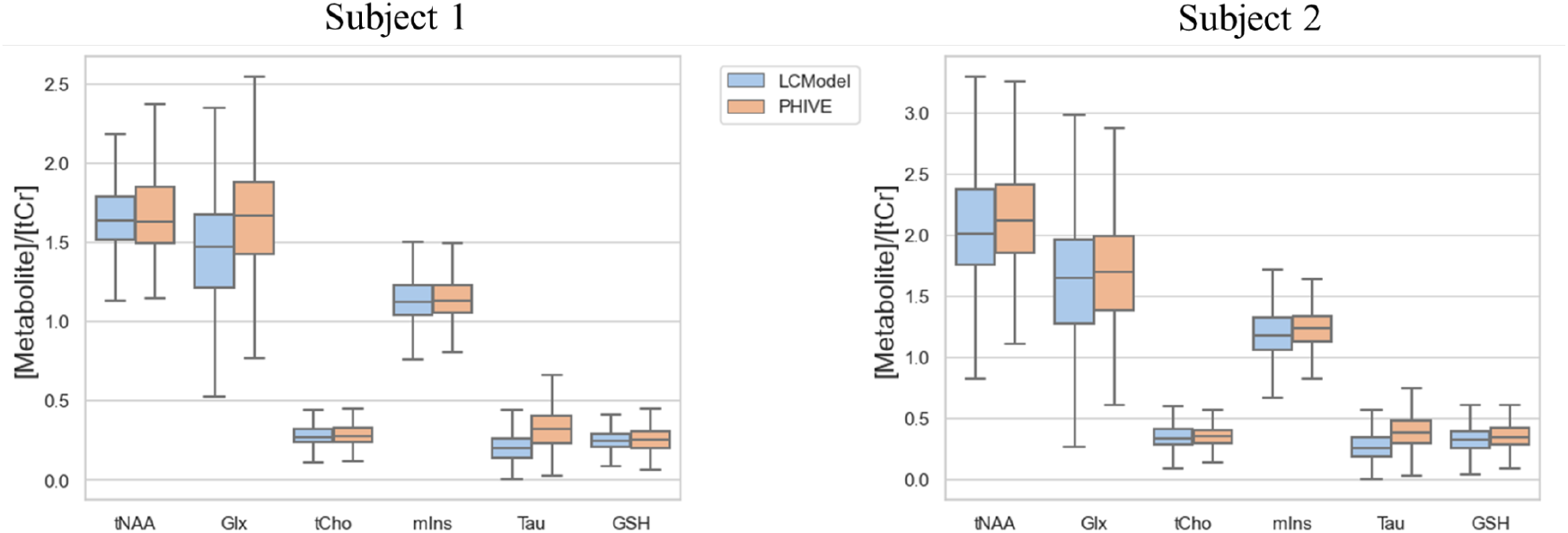
Comparison of metabolite relative concentrations obtained using LCModel and PHIVE normalized by tCr for six key metabolite ratios: tNAA, Glx, tCho, mIns, Tau, and GSH. The box plots display the median, interquartile range, and minimum/maximum values of the relative concentrations.

Figure 5 illustrates the spectral alignment and quantification results obtained using LCModel and our proposed PHIVE method. The brain MRI image on the left shows the region of interest from which the spectra were acquired. The plot on the right displays the overlaid spectra processed by LCModel (blue) and PHIVE (orange and green). It is important to note that LCModel applies a preprocessing step before quantification, which includes frequency correction. As a result, all spectra processed by LCModel are aligned in terms of their frequency. This preprocessing step ensures that the peaks of the metabolites are properly aligned across different spectra, facilitating accurate quantification. On the other hand, PHIVE does not include a separate preprocessing step for frequency correction. The PHIVE spectra shown in the plot exhibit variations in frequency alignment compared to the LCModel spectra. Despite these variations, the overall shape and peak positions of the PHIVE spectra closely resemble those of the LCModel spectra.

**Fig. 5.**
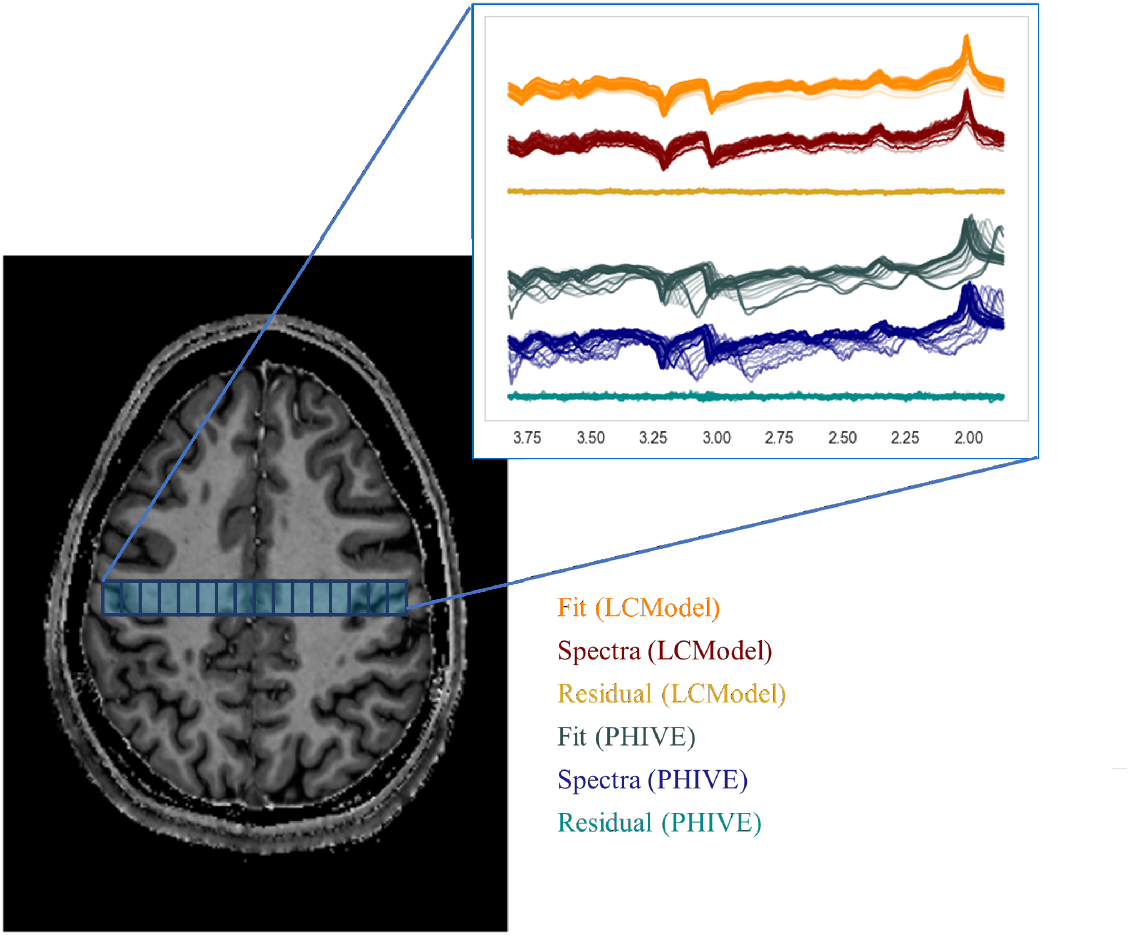
Comparison of spectral alignment and quantification results obtained using LCModel and PHIVE. The brain MRI image on the left shows the region of interest for spectral acquisition. The plot on the right displays the overlaid spectra processed by LCModel (blue), PHIVE with deep learning only (orange), and PHIVE with deep learning and basis set (green). LCModel applies a preprocessing step for frequency correction, resulting in aligned spectra, while PHIVE spectra exhibit slight variations in frequency alignment. The vertical dotted lines indicate the expected peak positions of key metabolites.

In figure 6, the spectra illustrate the comparison of spectral fitting results at varying SNR using the PHIVE model. The displayed spectra include the original signal, the fitted model, the quantified macromolecular spectrum, the spline baseline, and the residuals. Across different SNR levels, the fitting accuracy and residuals highlight the robustness of PHIVE in handling low-SNR data. At higher SNRs, the residuals are minimal, demonstrating accurate metabolite quantification and effective baseline subtraction. In contrast, at lower SNRs, while residuals are more pronounced, the model maintains a coherent spectral fit, capturing the key features of the data.

**Fig. 6.**
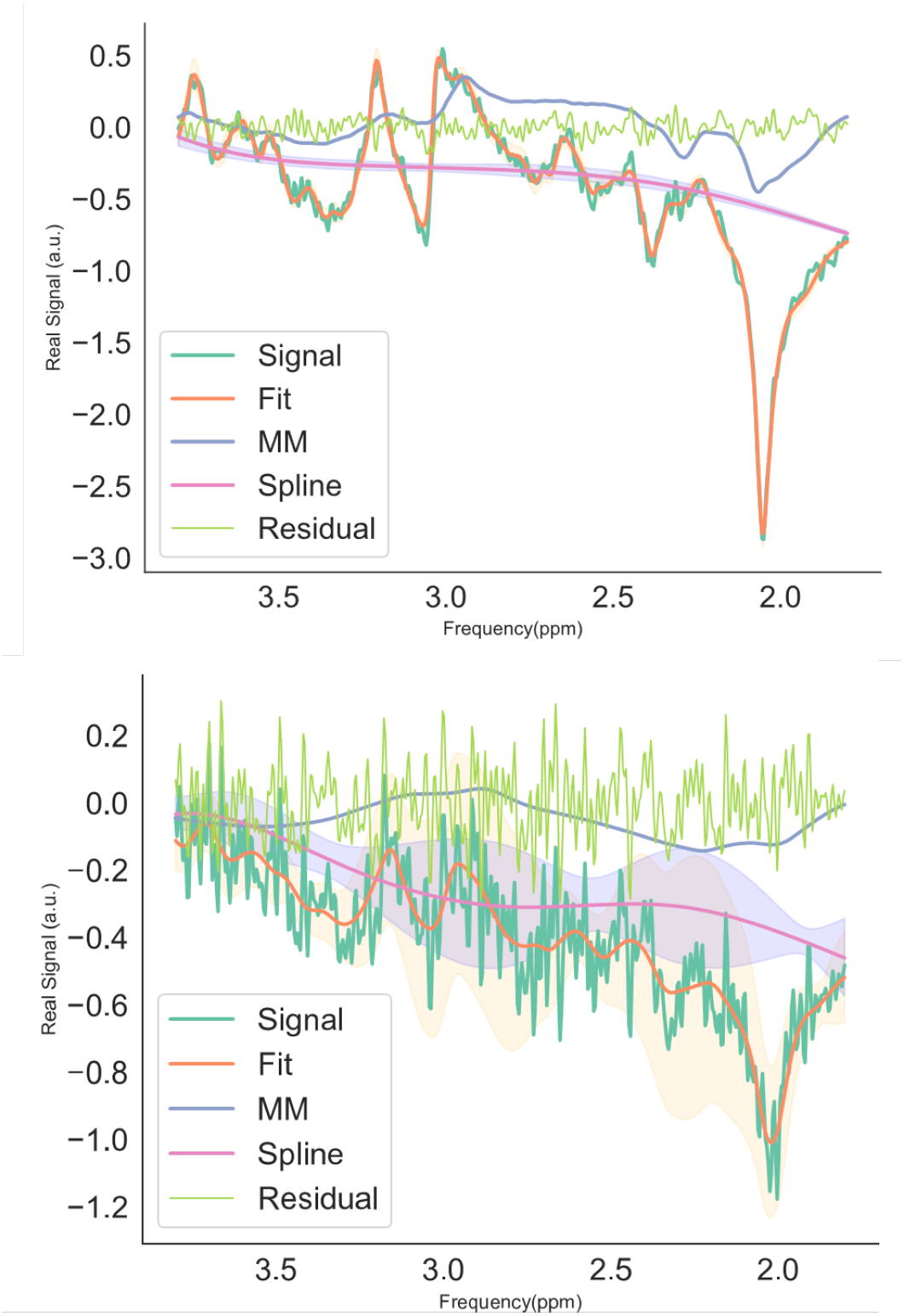
Sample spectra from a test subject along with the respective spectral fits, quantified macromolecular spectra, spline baseline and residual

Figure 7 presents the metabolite relative concentration maps and associated uncertainty estimates generated by the trained PHIVE model for a representative test subject. The model quantified the relative relative concentrations of tNAA, Glx, tCho, and mIns, providing spatial maps of their distributions normalized by tCr. Alongside the metabolite maps, PHIVE generates uncertainty estimates in the form of CRLB, epistemic uncertainty, and aleatoric uncertainty maps. The CRLB maps provide a lower bound on the variance of the estimated metabolite relative concentrations, with higher values indicating greater uncertainty. The epistemic uncertainty maps capture the model’s lack of knowledge about the optimal parameters, while the aleatoric uncertainty maps represent the inherent noise in the data. Across all metabolites, the uncertainty estimates are generally higher in regions with lower SNR, such as in the periphery and ventricles. The epistemic uncertainty maps exhibit more spatial variability compared to the aleatoric uncertainty maps, suggesting that the model’s parameter uncertainty varies across different brain regions. The CRLB maps show similar patterns to the epistemic uncertainty maps, although with less spatial heterogeneity.

**Fig. 7.**
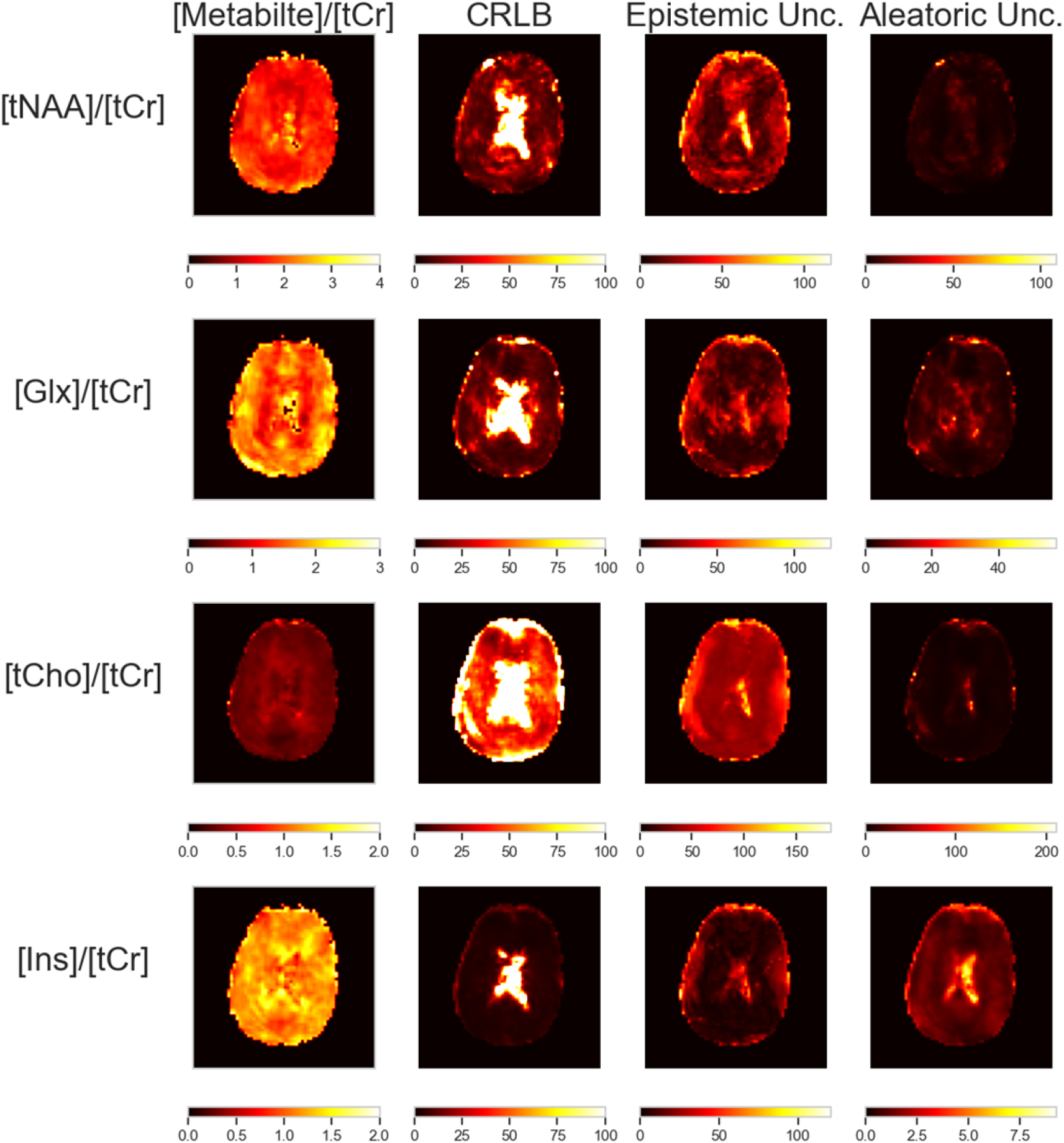
Metabolite relative concentration maps and associated uncertainty estimates generated by the PHIVE model. Each row corresponds to a different metabolite: tNAA/tCr, Glx/tCr, tCho/tCr, and mIns/tCr. The first column shows the metabolite relative concentration maps normalized by tCr. The remaining columns represent the CRLB, epistemic uncertainty, and aleatoric uncertainty maps, respectively. Higher intensity values in the uncertainty maps indicate greater uncertainty in the corresponding regions of the metabolite relative concentration maps.

Figure 8 shows a comparison of metabolite ratio maps between LCModel and PHIVE with varying K values (3, 6, 9, and 12). The figure shows the spatial distribution of four metabolite ratios: tNAA/tCr, Glx/tCr, tCho/tCr, and mIns/tCr. For each ratio, the LCModel result is presented alongside PHIVE results for different K values, with corresponding difference maps. The PHIVE method, employing our proposed conditional baseline approach, demonstrates varying performance across K values and metabolites. For tNAA/tCr and Glx/tCr, the difference maps reveal increasing spatial heterogeneity as K increases, with more pronounced differences in cortical regions at higher K values. This suggests that the baseline flexibility, controlled by the K parameter, significantly impacts these metabolite quantifications. tCho/tCr maps show relatively consistent differences across K values, indicating less sensitivity to changes in baseline flexibility for this metabolite ratio. However, subtle variations in difference patterns are observable, particularly in posterior regions. mIns/tCr exhibits the most dramatic changes with increasing K. At K=3, differences are relatively uniform, but as K increases to 12, more localized and intense differences emerge, especially in central and posterior brain regions.

**Fig. 8.**
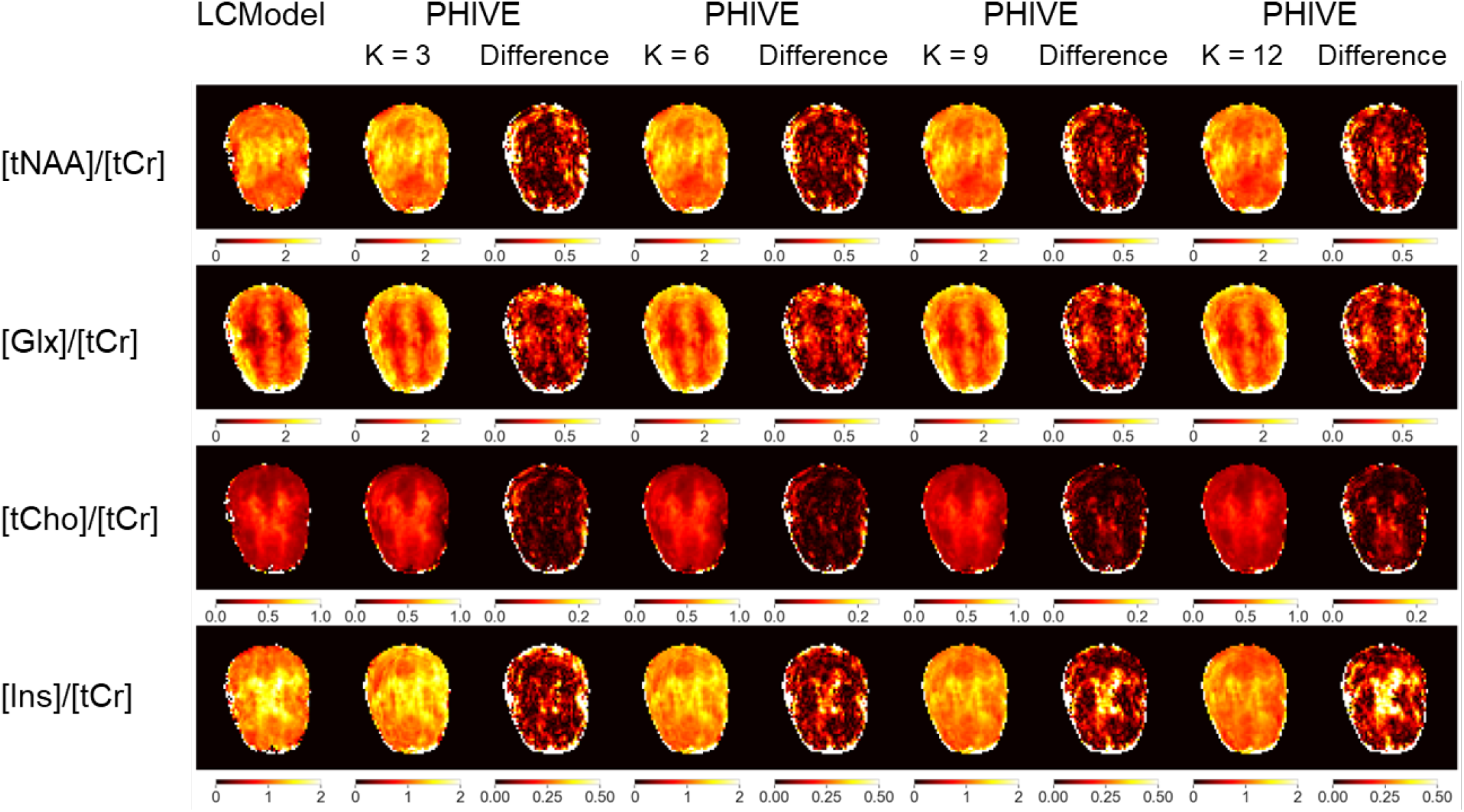
Visualization of metabolite quantification results across varying baseline stiffness levels. Each row represents a different metabolite, and columns illustrate the impact of conditioning on different baseline stiffness levels, ranging from 0 (most flexible) to 4 (most rigid). The heatmaps display metabolite relative concentration estimates, highlighting how varying baseline flexibility influences the accuracy of metabolite quantification. This approach dynamically adjusts the baseline based on input stiffness levels, thereby improving adaptability and efficiency without the need for multiple models.

## 4.1. Discussion

Our study introduces PHIVE, a novel deep learning framework for rapid spectral fitting in high-resolution whole-brain MRSI with simultaneous uncertainty estimation. The results demonstrate that PHIVE can achieve comparable accuracy to the gold standard LCModel while offering significant improvements in processing speed and providing comprehensive uncertainty quantification.

### 4.2. Performance and Accuracy

PHIVE demonstrated strong agreement with LCModel across multiple metabolites, particularly for NAA and Glx. The high correlation coefficients (*R*^2^ ranging from 0.49 to 0.82) and the visual similarity of the metabolite maps indicate that PHIVE can reliably quantify key brain metabolites. The slightly lower correlations for tCho and mIns (*R*^2^ = 0.49 and 0.50, respectively) suggest that these metabolites may be more challenging to quantify, possibly due to their lower relative concentrations or spectral overlap with other metabolites.

Importantly, PHIVE consistently demonstrated lower CVs compared to LCModel for most metabolites across all subjects. This improved precision is particularly notable for Glx and mIns, where PHIVE achieved CVs ranging from 22.10% to 24.10% and 11.15% to 13.47%, respectively, compared to LCModel’s 24.17% to 28.70% and 15.28% to 16.02%. The enhanced precision of PHIVE could be attributed to its ability to leverage spatial information and learn complex patterns from the entire dataset, potentially reducing the impact of noise and artifacts on individual voxel quantification.

### 4.3. Computational Efficiency

One of the most striking advantages of PHIVE is its computational efficiency. The testing phase took only 6 milliseconds per subject, representing a reduction in computational time by six orders of magnitude compared to the conventional LCModel approach. This dramatic speedup has significant implications for clinical applications, potentially enabling real-time metabolite quantification and integration into clinical workflows.

### 4.4. Uncertainty Quantification

A key innovation of PHIVE is its ability to provide comprehensive uncertainty estimates, including CRLB, epistemic uncertainty, and aleatoric uncertainty. The uncertainty maps reveal higher uncertainty in regions with lower SNR, such as the brain periphery and ventricles, which aligns with our understanding of MRSI data quality. The distinction between epistemic and aleatoric uncertainty offers valuable insights into the sources of uncertainty, with epistemic uncertainty showing more spatial variability. This detailed uncertainty quantification could enhance the interpretability and reliability of MRSI results in both research and clinical settings.

### 4.5. Implications of Conditional Baseline Modeling

The varying patterns observed in the difference maps across K values align with our conditional baseline approach, demonstrating the method’s capability to capture different aspects of metabolite distributions based on the chosen stiffness level. This flexibility in adjusting baseline stiffness through the K parameter offers a nuanced approach to metabolite quantification, potentially allowing for optimization of the baseline model for specific metabolites or brain regions. However, it is important to note that while our method provides this flexibility, the optimal choice of baseline stiffness remains an open area of research. The complex interplay between baseline modeling and metabolite quantification accuracy necessitates further investigation. Future studies should focus on developing systematic approaches to determine the most appropriate stiffness level for different metabolites and brain regions, considering factors such as SNR, spectral quality, and specific research or clinical objectives. This continued research will be crucial in fully leveraging the potential of flexible baseline modeling in MRSI.

### 4.6. Limitations and Future Directions

Despite the promising results, several limitations and areas for future research should be addressed:

1. **Lack of ground truth:** As with most MRSI studies, the absence of a true gold standard for in vivo metabolite relative concentrations limits our ability to assess absolute accuracy. Future work could involve phantom studies or simulations with known metabolite relative concentrations to further validate PHIVE’s performance.
2. **Limited sample size:** Our study included a relatively small number of subjects. Larger, multi-center studies would be beneficial to assess the generalizability of PHIVE across different scanners and patient populations.
3. **Spectral preprocessing:** Unlike LCModel, PHIVE does not include a separate preprocessing step for frequency correction. While this simplifies the pipeline, it may impact the accuracy of quantification, especially in regions with large B0 inhomogeneities. Future iterations of PHIVE could incorporate automated preprocessing steps to address this limitation.
4. **Metabolite specificity:** The lower correlations observed for tCho and mIns suggest that PHIVE may have difficulty distinguishing these metabolites. Further refinement of the model architecture or incorporation of prior knowledge about metabolite ratios could potentially improve the specificity of quantification.
5. **Interpretability:** Despite the physics-informed nature of PHIVE, the deep learning approach may be perceived as a ”black box” by some clinicians. Developing methods to enhance the interpretability of the model’s decisions could increase its acceptance in clinical practice.
6. **Longitudinal studies:** Investigating PHIVE’s performance in longitudinal studies would be valuable to assess its sensitivity to metabolic changes over time and its potential for monitoring disease progression or treatment response.

### 4.7. Conclusion

In conclusion, PHIVE represents a significant advancement in MRSI analysis, offering rapid, accurate, and uncertainty-aware metabolite quantification. Its ability to process wholebrain MRSI data in milliseconds while providing comprehensive uncertainty estimates has the potential to transform the use of MRSI in both research and clinical settings. Future work should focus on addressing the identified limitations and validating PHIVE across diverse datasets and clinical applications. As deep learning continues to evolve, frameworks like PHIVE are likely to play an increasingly important role in extracting meaningful information from complex neuroimaging data, ultimately contributing to improved understanding and management of neurological disorders.

## Data Availability

All data produced in the present study are available upon reasonable request to the authors

## Notes

### Competing Interest Statement

The authors have declared no competing interest.

### Funding Statement

This project has received funding from the European Union's Horizon 2020 research and innovation program under the Marie Skłodowska-Curie grant agreement No 813120, and Austrian Research Fund P 34198.

### Author Declarations

This study was performed in line with the principles of the Declaration of Helsinki. Approval was granted by the Ethics Committee of Medical University of vienna EC 2009/154.

